# Neuromelanin-sensitive MRI correlates of cognitive and motor function in Parkinson’s disease with freezing of gait

**DOI:** 10.1101/2023.07.04.23292227

**Authors:** Daniel E. Huddleston, Xiangchuan Chen, Kristy Hwang, Jason Langley, Richa Tripathi, Kelsey Tucker, J. Lucas McKay, Xiaoping Hu, Stewart A. Factor

## Abstract

Substantia nigra pars compacta (SNc) and locus coeruleus (LC) are neuromelanin-rich nuclei implicated in diverse cognitive and motor processes in normal brain function and disease. However, their roles in aging and neurodegenerative disease mechanisms have remained unclear due to a lack of tools to study them *in vivo*. Preclinical and post-mortem human investigations indicate that the relationship between tissue neuromelanin content and neurodegeneration is complex. Neuromelanin exhibits both neuroprotective and cytotoxic characteristics, and tissue neuromelanin content varies across the lifespan, exhibiting an inverted U-shaped relationship with age. Neuromelanin-sensitive MRI (NM-MRI) is an emerging modality that allows measurement of neuromelanin-associated contrast in SNc and LC in humans. NM-MRI robustly detects disease effects in these structures in neurodegenerative and psychiatric conditions, including Parkinson’s disease (PD). Previous NM-MRI studies of PD have largely focused on detecting disease group effects, but few studies have reported NM-MRI correlations with phenotype. Because neuromelanin dynamics are complex, we hypothesize that they are best interpreted in the context of both disease stage and aging, with neuromelanin loss correlating with symptoms most clearly in advanced stages where neuromelanin loss and neurodegeneration are coupled. We tested this hypothesis using NM-MRI to measure SNc and LC volumes in healthy older adult control individuals and in PD patients with and without freezing of gait (FOG), a severe and disabling PD symptom. We assessed for group differences and correlations between NM-MRI measures and aging, cognition and motor deficits. SNc volume was significantly decreased in PD with FOG compared to controls. SNc volume correlated significantly with motor symptoms and cognitive measures in PD with FOG, but not in PD without FOG. SNc volume correlated significantly with aging in PD. When PD patients were stratified by disease duration, SNc volume correlated with aging, cognition, and motor deficits only in PD with disease duration >5 years. We conclude that in severe or advanced PD, identified by either FOG or disease duration >5 years, the observed correlations between SNc volume and aging, cognition, and motor function may reflect the coupling of neuromelanin loss with neurodegeneration and the associated emergence of a linear relationship between NM-MRI measures and phenotype.

## Introduction

The neuromelanin-rich catecholamine nuclei, substantia nigra pars compacta (SNc) and locus coeruleus (LC), are implicated in diverse cognitive and motor processes in normal physiology and disease states. However, *in vivo* study of their roles in human brain function has remained difficult due to a lack of adequate tools. It has also been challenging to clarify the mechanistic role of neuromelanin in Parkinson’s disease (PD) because its roles in normal cellular functioning and in disease states are complex. Neuromelanin exhibits both neuroprotective and cytotoxic characteristics in SNc and LC neurons, and is impacted differently by the aging process and neurodegenerative disease.^1^ In humans neuromelanin pigment first appears at about age three and it accumulates with age. ^2–7^ Neuromelanin formation is itself a neuroprotective process, which involves removal of neurotoxic substances from the cytosol.^8, 9^ Its synthesis is driven by excess cytosolic dopamine in catecholamine neurons, and it sequesters iron, toxic dopamine metabolites (e.g. dopamine-o-quinone), and a mixture of aggregated proteins, including α-synuclein.^9, 10^ By sequestering these substances neuromelanin synthesis prevents neurodegeneration.^1^

The accumulation of neuromelanin with age does not continue indefinitely, however, and neuromelanin content in SNc and LC peaks in late middle age, between about age 50 and 60, and then declines.^11–14^ Consequently, over the human lifespan SNc and LC neuromelanin content follows an inverted U-shaped curve.^11, 14^ In PD and related forms of neurodegenerative parkinsonism selective loss of melanized neurons occurs in these structures, but the mechanistic role of neuromelanin in these conditions has not been clarified.^13, 15–17^ The early accumulation of neuromelanin during periods of normal brain function, which is then followed by a decline that occurs in parallel with neurodegeneration implies a possible role for neuromelanin in PD pathogenesis. Exploration of this hypothesis in animal models has been limited, though, because rodents do not produce neuromelanin, and therefore rodent models of PD have not included study of its biology. This has changed, recently, with the development of a new rodent model of PD that produces neuromelanin in SNc.^18^ Work using this model demonstrates that after a threshold concentration is reached, gradually increasing levels of intracellular neuromelanin are associated with cell sickness, with an initial loss of protein expression followed by nigrostriatal neurodegeneration.^19, 20^ The model develops Lewy body-like inclusions, and a hypokinetic parkinsonian phenotype. Furthermore, the phenotype can be rescued with an over-expression of vesicular monoamine transporter 2 (VMAT2), which causes increased sequestration of cytosolic dopamine, thereby decreasing neuromelanin-synthesis.^21^ Other research has shown that neurodegeneration-associated release of neuromelanin granules into the extracellular space activates microglia and drives neuroinflammation.^8, 22, 23^ Clearance of neuromelanin from SNc in advanced PD is a long-established finding on gross pathology, and postmortem studies suggest that activated microglia may clear extracellular neuromelanin, which accumulates secondary to neuronal death.^23–25^ The complex dynamics of neuromelanin in the biology of aging and neurodegenerative disease are depicted schematically in **Figure 1**. In combination, these insights suggest that *in vivo* monitoring of tissue neuromelanin content may yield insights into the biology of parkinsonian neurodegeneration, and modulation of neuromelanin biology may be a viable therapeutic strategy.

**Figure 1.**
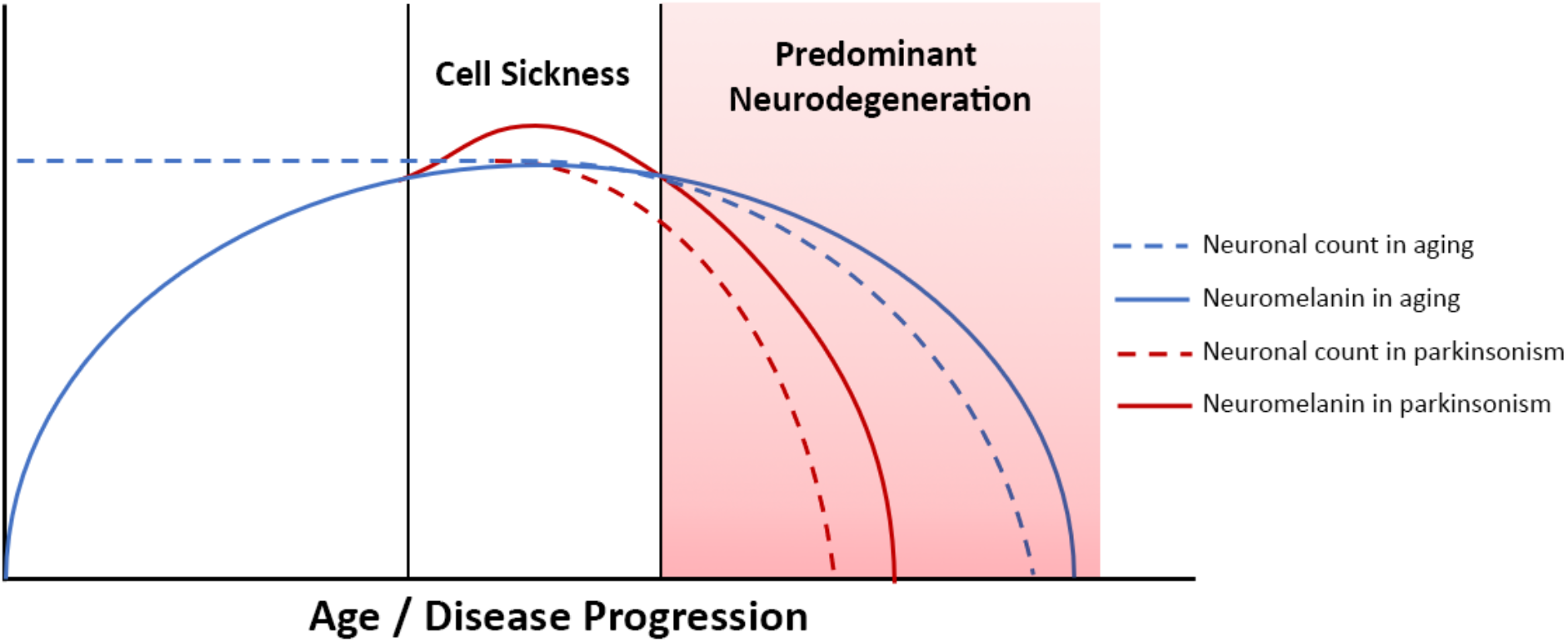
Neuromelanin dynamics in aging and neurodegenerative parkinsonism. Advanced PD is associated with neurodegeneration and neuromelanin loss, whereas in early PD tissue neuromelanin content and neurodegeneration are not yet coupled. Correlation between phenotypic progression and neuromelanin loss are therefore most easily observed in more severe/advanced disease.

Neuromelanin-sensitive MRI (NM-MRI) measurement of SNc and LC volumes has emerged as a highly reproducible modality to measure tissue neuromelanin *in vivo* and to detect disease effects.^26, 27^ NM-MRI contrast in these structures is a result of magnetization transfer (MT) effects, and both explicit and incidental MT contrast approaches have been used.^24^ In this study an MT prepared gradient echo (MT-GRE) NM-MRI pulse sequence was used because this approach has previously been shown to exhibit improved contrast-to-noise ratio (CNR) in SNc imaging as compared to incidental MT-based NM-MRI sequences.^28^ Both the MT-GRE sequence and the automated image processing method for threshold-based volume measurement of LC and SNc applied here have been shown to have superior scan-rescan reproducibility compared to incidental MT-based NM-MRI and manual segmentation approaches for SNc and LC measurement.^27, 29, 30^ These methods were chosen because highly reproducible NM-MRI methods with favorable CNR are necessary to advance our understanding of neuromelanin biology and to identify candidate biomarkers for translational applications.

In this study we applied these tools to study PD with and without freezing of gait (FOG). FOG is a common and disabling feature of PD. FOG is a leading cause of falls and immobility, and it leads to social isolation and reduced quality of life.^31–36^ Its prevalence increases with disease progression; at 10 years of disease duration approximately 60-70% of PD patients exhibit FOG.^37, 38^ Thus, FOG may be considered a marker of advanced motor dysfunction in PD. Both dopaminergic and noradrenergic dysfunction are implicated in FOG.^39, 40^ However, so far no studies have reported NM-MRI assessment of SNc and LC volumes in PD patients whose FOG status is characterized, nor have any studies assessed NM-MRI correlation with cognitive and motor phenotype in early vs. advanced PD stages.

Here we report our investigation of LC and SNc volumes in healthy older adults, PD without FOG (PD-NoFOG) and PD with FOG (PD-FOG) using NM-MRI. We assess for correlations between NM-MRI measurements of LC and SNc volumes and cognitive and motor aspects of PD. Specifically, we investigate relations between these widely projecting nuclei and global cognition, cognitive reserve, parkinsonian motor dysfunction, FOG, and aging in PD. The objectives of the study were to test in living humans the hypotheses implied in **Figure 1**: A) in advanced PD a linear decline in neuromelanin is associated with neurodegeneration and the motor and cognitive phenotypes associated with neurodegeneration, and B) increasing age exacerbates neurodegeneration in LC and SNc. To address these questions in a cross-sectional study design, in one set of analyses the PD groups were stratified based on the presence vs. absence of FOG, a symptom of severe motor dysfunction. This is useful because rates of disease progression can vary widely between patients with PD, and FOG can identify clinically advanced PD independent of disease duration. Correlations were then assessed between NM-MRI derived SNc volumes and measures of motor and cognitive function in early vs. advanced PD, defined by the presence or absence of FOG. However, because disease duration may capture aspects of PD biological progression differently than FOG status, a second set of analyses were performed in early vs. advanced PD, defined by shorter (< 5 years) vs. longer (≥ 5 years) disease duration. Finally, we assessed for correlations between age and LC volume or SNc volume in PD-FOG, PD-NoFOG, PD with < 5 years duration, PD with ≥ 5 years duration, and older adult control individuals.

We hypothesized that linear relationships between LC and SNc integrity and motor and cognitive disease features will be most readily observed in more advanced PD, defined by the presence of the more severe motor phenotype, FOG, or based on longer disease duration, i.e., > 5 years. This was expected because neurodegenerative effects on tissue neuromelanin content and NM-MRI contrast will predominate in advancing disease. This, in turn, is expected to cause the emergence of linear associations between NM-MRI and neurodegeneration-associated motor and cognitive phenotypes.

Conversely, NM-MRI contrast is expected to plateau earlier in the disease course due to the competing effects of cell sickness (neuromelanin accumulation) and neurodegeneration (neuromelanin loss). As a result of these complex effects influencing NM-MRI contrast, we do not expect to observe linear relationships between NM-MRI and neurodegeneration-associated phenotypes in early disease. This hypothesis is based on the understanding of neuromelanin dynamics in aging and neurodegenerative disease reviewed above and displayed schematically in **Figure 1**.

## Methods

### Research participants

Forty-nine individuals with PD meeting the UK Brain Bank Criteria as assessed by a fellowship-trained movement disorders neurologist were recruited from the Emory Movement Disorders Clinic.^41^ Nineteen older adult control participants were recruited from the Emory Alzheimer’s Disease Research Center. FOG status was determined by either 1) a combination of clinical assessment by a movement disorder neurologist and Freezing of Gait Questionnaire (FOGQ) item 3 score ;: 2, or 2) a structured clinical examination in the practically-defined OFF state and after a levodopa challenge as previously reported.^39, 42^ Mean dose of levodopa challenge was approximately 140% of the patient’s typical dose. Participants in the Emory Freezing of Gait Study were assessed by a movement disorders neurologist for the presence or absence of FOG in three conditions: 1) 360 degree turns to left (twice) and right (twice), 2) timed up-and-go testing consisting of walking 10 feet, turning and returning to the starting position, and 3) the same timed up-and-go task done while performing serial calculations. Of the 49 PD participants, 19 were found to have no FOG and 30 were found to have FOG.

All study participants were clinically assessed with the UPDRS-III or the MDS-UPDRS-III as well as the Montreal Cognitive Assessment (MoCA).^43–45^ UPDRS-III scores were converted to MDS-UPDRS-III scores using the conversion approach published by Hentz and colleagues, and MDS-UPDRS-III scores were used for analyses.^46^

### Image acquisition and processing

Each research participant underwent MRI scanning using one of two Siemens Trio 3 T scanners using a 12-channel receive only head coil at the Emory Center for Systems Imaging. NM-MRI data was acquired using a 2D GRE pulse sequence with a reduced flip angle MT preparation pulse with additional parameters as previously published.^47^ Image processing steps to determine SNc volume and LC volume were carried out using an automated, published approach, using FMRIB Software Library (FSL) and custom scripts developed in MATLAB (The Mathworks, Natick, MA).^27^ Due to rare errors in field of view placement at the time of acquisition, SNc volume could not be calculated for one participant who had PD-FOG, and LC volume could not be calculated on a different individual with PD-FOG.

### Statistical Analysis

Differences in central tendency between study groups were assessed with ANOVA or Chi square tests as appropriate. Analyses of covariance (ANCOVA) were used to compare SNc volumes and LC volumes between groups while controlling for age, sex, years of education and scanner type. Pairwise comparisons following ANCOVA were performed using Fisher’s least significant difference. Statistical analyses were performed in SPSS 26.0 (IBM) and plots were generated using GraphPad Prism 9.0.0.

Stratified analyses assessed relationships between SNc volume and motor and cognitive outcomes separately for PD patients with and without FOG as well as for all PD patients in the sample. One PD-FOG patient did not have an available SNc volume and another PD-FOG patient did not have an available LC volume value. Pearson correlation coefficients were calculated between SNc volume and MoCA, education, MDS-UPDRS-III, and FOGQ score. Considering the potential for ceiling effects with the MoCA and floor effects with the FOGQ score that could cause departures from normality, we used Spearman correlation as appropriate when the data were not normally distributed. Identified correlation coefficients were compared with partial correlation coefficients controlling for disease duration to determine whether imbalances in PD duration could impact significant findings. Additional correlation analyses were performed stratified on disease duration ≤5 or >5 years. One-tailed correlations were used in all correlational analyses because the direction of the predicted effect is clearly specified (lower volume in SNc or LC, consistent with neurodegeneration, is expected to correlate with worsened symptoms, i.e., higher MDS-UPDRS-III, lower MoCA, etc.).

## Results

Clinical and demographic characteristics of the study groups are shown in **Table 1**. Healthy controls were significantly older (71 vs. 62 and 67 years) and had additional years of education (18 vs. 16 years) compared to the two PD groups (**Table 1**). Compared to PD-NoFOG, PD-FOG patients had significantly longer disease duration (9.6 vs. 5.8 years), higher levodopa equivalents (1333 vs. 695 mg), and higher FOGQ scores (17.3 vs. 3.7) There were no significant differences in MDS-UPDRS-III scores between PD with and without FOG (p = 0.172).

**Table 1.** Demographic and clinical data.

| **Group characteristics**<br>**(Mean ± Standard Error)** | **Healthy Control**<br>**(N=19)** | **PD-NoFOG**<br>**(N=19)** | **PD-FOG**<br>**(N=30)** | **p-value** |
| --- | --- | --- | --- | --- |
| **Age** | 71.3 ± 1.2 | 62.1 ± 2.7 | 67.3 ± 1.4 | **0.005** |
| **Gender (M:F)** | 9:10 | 13:6 | 24:6 | 0.059 |
| **Education (years)** | 18.2 ± 0.5 | 16.0 ± 0.5 | 16.6 ± 0.5 | **0.014** |
| **Disease Duration (years)** | - | 5.8 ± 0.7 | 9.6 ± 1.2<br>(N=29) | **0.007** |
| **MDS-UPDRS-III ON medication** | - | 28.9 ± 3.2 | 25.3 ± 2.4 | 0.361 |
| **LEDD** | - | 694.6 ± 66.8 | 1332.8 ± 142.4 | **2.2×10<sup>-4</sup>** |
| **MoCA** | 27.5 ± 0.5 | 26.5 ± 0.9 | 24.6 ± 0.9 | **0.044** |
| **FOGQ** | - | 3.7 ± 1.2 | 17.3 ± 1.1 | **2.7×10<sup>-10</sup>** |

**LEDD: levodopa equivalent daily dose**

Representative NM-MRI images of SNc of controls and PD patients with and without FOG are shown in **Figure 2**. Mean SNc volume varied significantly across groups (p = 0.019, ANCOVA) after controlling for age, gender, years of education, and scanner type. Mean SNc volumes are shown in **Figure 3** (mean ± SE: control = 448.9 ± 30.7, PD-NoFOG = 373.5 ± 25.2, PD-FOG = 330.1 ± 21.0).

**Figure 2.**
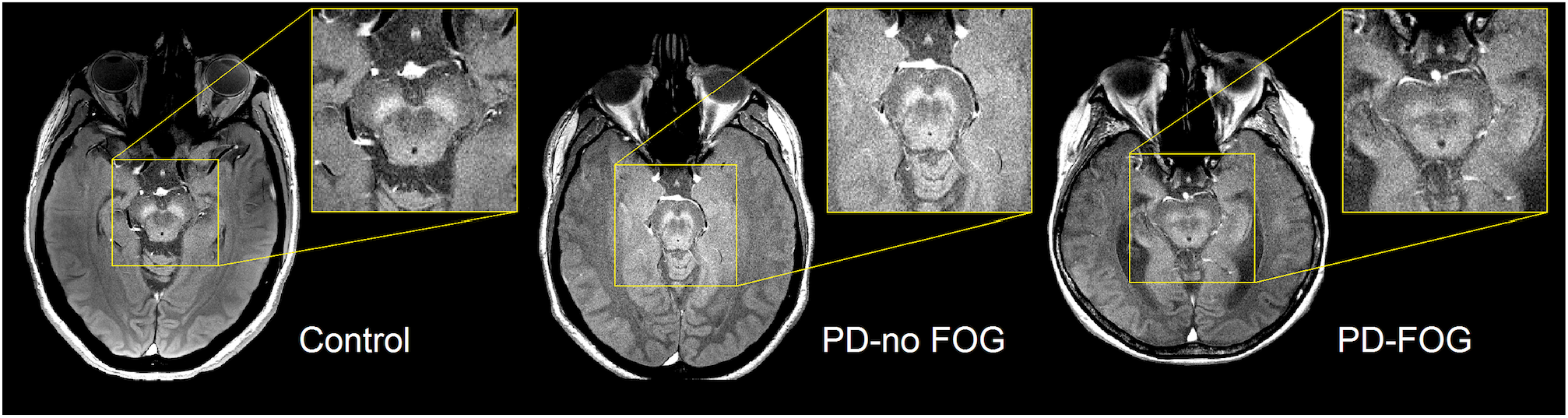
Representative images of NM-MRI contrast in SNc of healthy controls and PD patients with or without FOG. Hyperintense contrast in SNc of a healthy control individual acquired by NM-sensitive MRI. SNc NM-MRI signal intensity is decreased in PD-NoFOG compared to controls, and an even more profound loss of hyperintensity is seen in PD-FOG.

**Figure 3.**
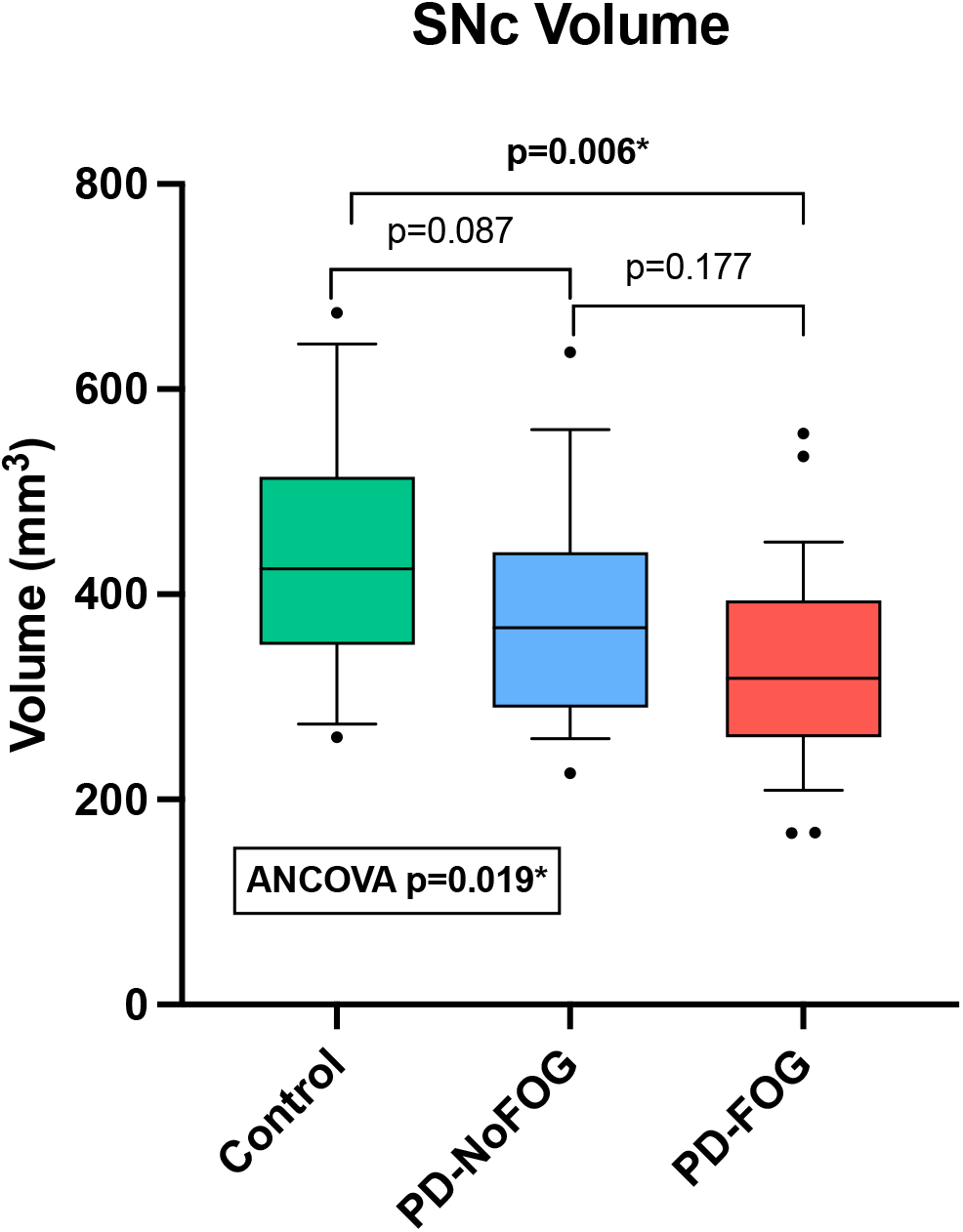
Comparison of SNc volumes between healthy controls, PD-NoFOG, and PD-FOG. ANCOVA including age, gender, years of education, and scanner type as covariates. Control N=19, PD-NoFOG N=19, PD-FOG N=29. Boxes represent 25^th^ to 75^th^ percentiles, whiskers show 10^th^ to 90^th^ percentiles. Line is median. * p<0.05.

Pairwise comparisons with Fisher’s least significant difference found a significant difference in SNc volume between the healthy controls and PD-FOG group (p = 0.006), but not between controls and the PD-NoFOG group (p = 0.087), nor between PD-NoFOG and PD-FOG (p = 0.177). No significant differences in mean LC volume across groups were identified using similar ANCOVA models controlling for age, gender, education, and scanner type (p = 0.073; mean ± SE: control = 7.34 ± 0.92, PD-NoFOG = 5.69 ± 0.76, PD-FOG = 4.50 ± 0.63).

### SNc correlations with cognitive and motor measures in PD, stratifying by FOG status

In PD-FOG, there were significant positive correlations between SNc volume and MoCA score (Pearson: r = 0.506, p = 0.003; spearman: rho = 0.498, p = 0.003) and between SNc volume and education (r = 0.355, p = 0.029), as shown in **Figure 4**. No significant correlations between these measures were observed in PD-NoFOG. Furthermore, similar relationships between SNc volume and MoCA or education were observed when the data were analyzed using models of partial correlation accounting for potential confounding effects of age and scanner type (SNc vs. MoCA: r = 0.394, p = 0.021; SNc vs. education: r = 0.345, p = 0.039).

**Figure 4.**
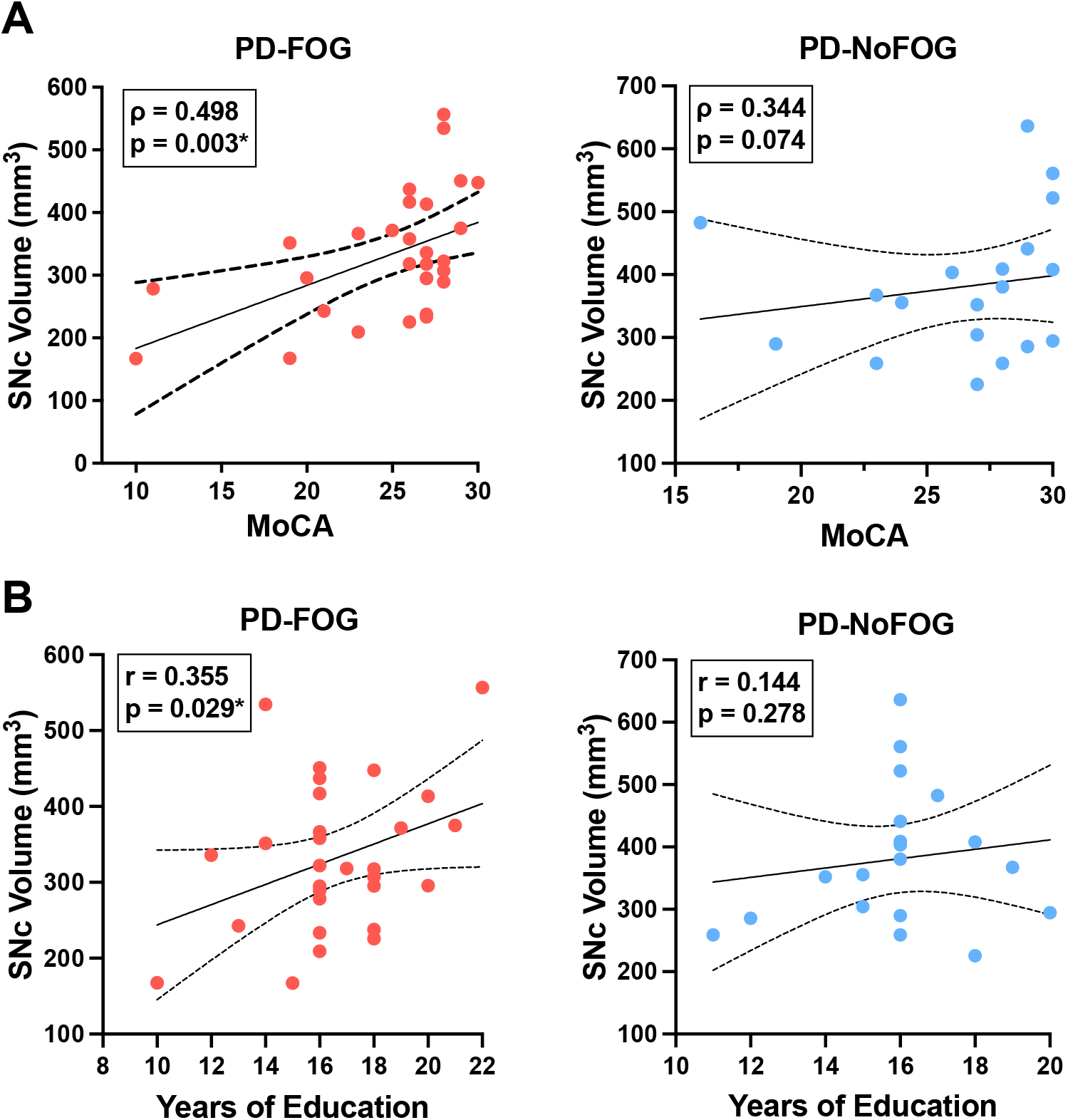
SNc volume vs. cognition and years of education in PD with and without FOG. SNc volume is positively correlated with MoCA (A, left) and years of education (B, left) in PD-FOG (N=29), but no significant correlation between these measures is seen in PD-NoFOG (A and B, right; N=19). Dotted lines represent 95% confidence intervals. * p<0.05.

SNc volume showed significant negative correlations with ON medications MDS-UPDRS-III (r = - 0.405, p = 0.015) and FOGQ scores (r = -0.318, p = 0.046) in the PD-FOG group, as shown in **Figure 5**. These relationships were also assessed using partial correlation controlling for age and scanner type, and the correlation between SNc volume and MDS-UPDRS-III remained significant (r = -0.328, p = 0.047), as did the correlation between SNc volume and FOGQ score (r = -0.401, p = 0.019). No significant correlations between SNc volume and ON medications MDS-UPDRS-III or FOGQ score were observed in the PD-NoFOG group. The relationship between SNc volume and FOGQ score in the PD-NoFOG group was also analyzed using Spearman’s correlation and remained non-significant (ρ = - 0.177, p = 0.234). Furthermore, this relationship remained non-significant following the omission from the analysis of the two apparent outliers with relatively high FOGQ scores (**Figure 5B, right**; ρ = -0.299, p = 0.124).

**Figure 5.**
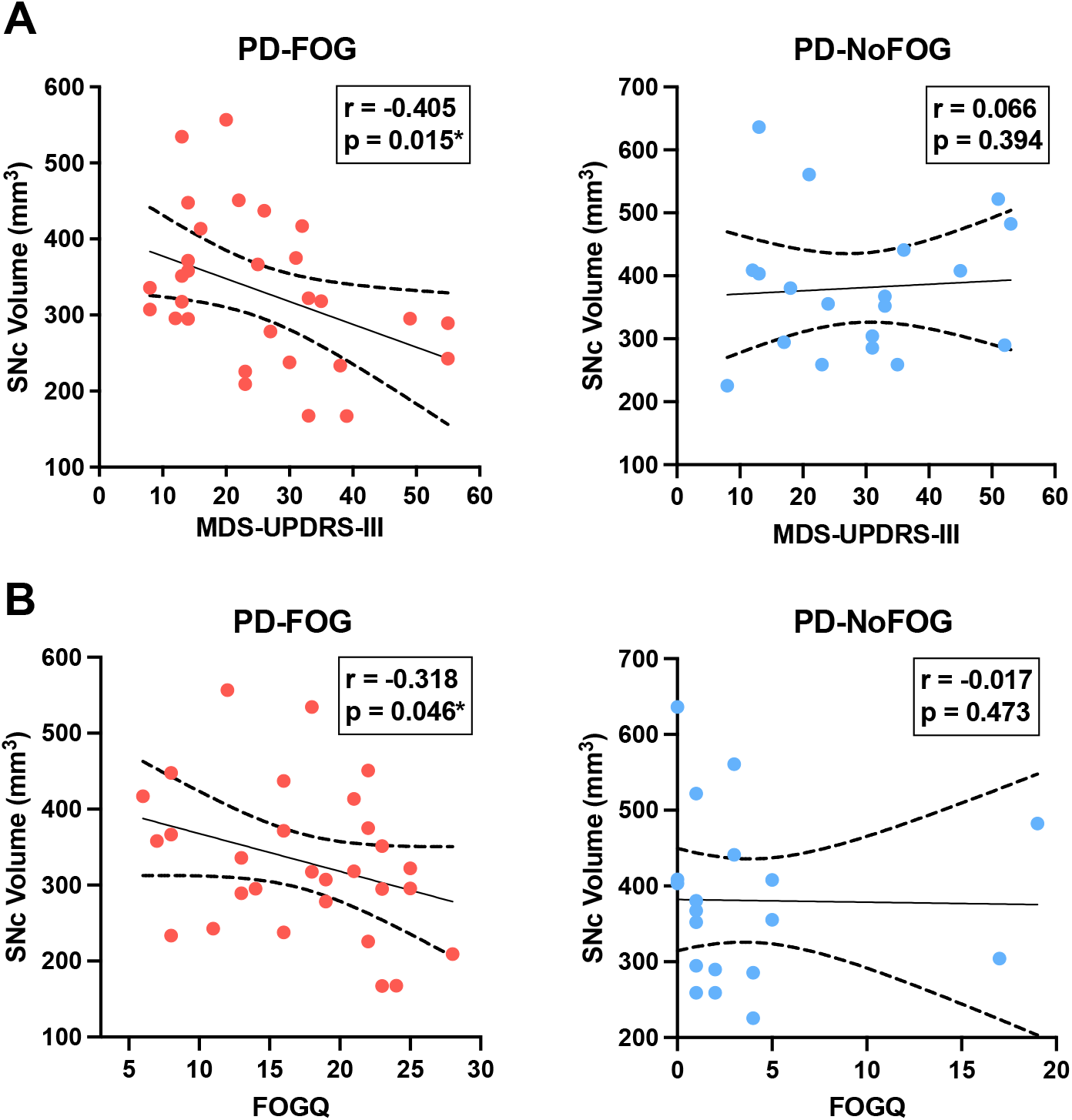
SNc volume vs. MDS-UPDRS-III motor score and FOGQ score in PD with and without FOG. SNc volume is negatively correlated with MDS-UPDRS-III (A, left) and FOGQ (B, left) in PD-FOG (N=29), but no significant correlation between these measures is observed in PD-NoFOG (A and B, right; N=19). Dotted lines represent 95% confidence intervals. *

### SNc correlations with cognitive and motor measures in PD, stratifying by disease duration

Among PD patients with > 5 years disease duration (N = 28), significant correlations were found between SNc volume and FOGQ score (r = -0.405, p = 0.016) and MoCA total score (r = 0.457, p = 0.007). Correlations between SNc volume and FOGQ score or MoCA remained significant when they were analyzed using partial correlation controlling for age and scanner type (p = 0.017, r = -0.410, SNc vs. FOGQ; p = 0.007, r = 0.457, SNc vs. MoCA). Among PD with ≤ 5 years disease duration (N = 19), no significant correlations were found between SNc volume and MDS-UPDRS-III score, FOGQ score, MoCA total score, years of education, or age.

### SNc and LC volume correlations with age in PD

SNc volume was strongly negatively correlated with age in the PD group (r = -0.462, p = 0.0005), while no correlation was observed between SNc volume and age in the older adult control group, as shown in **Figure 6A**. The correlation between SNc volume and age in the PD group remained significant (r = -0.477, p < 0.001) when controlling for scanner type in partial correlation analysis. Both LC and SNc volumes were significantly negatively associated with age in PD with disease duration > 5 years as shown in **Figure 6B** (LC: r = -0.436, p = 0.010; SNc: r = -0.633, p = 0.0002) and these correlations remained significant when controlling for scanner type in partial correlation analysis (LC: r = -0.442, p = 0.011; SNc: r = -0.629, p < 0.001). There were no significant correlations between LC volume and age or between SNc volume and age in PD with ≤ 5 years disease duration.

**Figure 6.**
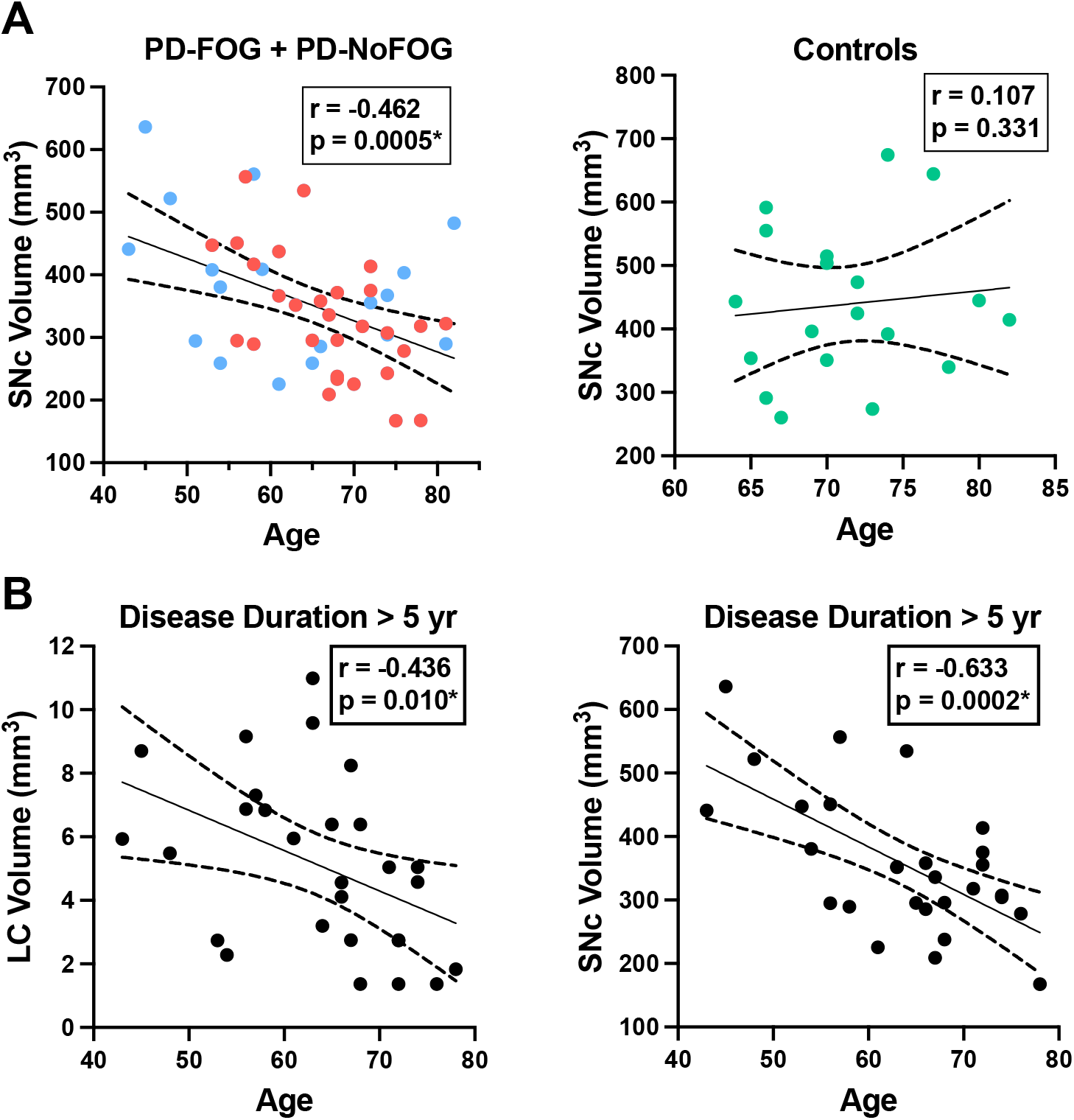
LC and SNc volumes vs. age in PD. **(A, left)** SNc volume exhibits a significant negative correlation with age in PD (N = 48, combined group with FOG shown in red (N=29) and without FOG shown in blue (N=19). **(A, right)** SNc volume is not correlated with age in older adult control participants (N = 19). **(B, left)** LC volume is negatively correlated with age in PD with disease duration > 5 years (N = 28). **(B, right)** SNc volume exhibits strong negative correlation with age in PD with disease duration > 5 years (N=28). Dotted lines

## Discussion

The baseline characteristics of the PD-NoFOG and PD-FOG groups in our study (**Table 1**) are consistent with other previous reports, as PD-FOG individuals were observed to have significantly higher levodopa equivalent daily dose (LEDD) than PD-NoFOG.^39, 48^ This was a strikingly large difference, as PD-FOG individuals were taking nearly double the LEDD as PD-NoFOG, despite the two groups having very similar MDS-UPDRS-III motor scores. This likely relates specifically to the limited levodopa response of FOG. The lack of difference in MDS-UPDRS-III score between the groups is important because this prevents overall motor severity from confounding our stratification of these groups based on FOG status. The longer disease duration observed in the PD-FOG group as compared to PD-NoFOG was also expected, because FOG is increasingly prevalent with longer disease duration in PD ^37, 38^. The trend towards worse global cognition found in PD-FOG as compared to PD-NoFOG and controls was also consistent with prior observations that FOG is associated with cognitive impairment ^49-52^.

Comparison of NM-MRI measurements of SNc volume between groups, shown in **Figure 3**, revealed that PD-FOG had significantly lower SNc volume than older adult controls. Numerically, PD-NoFOG had SNc volume that was intermediate between the control group and PD-FOG, although the differences between PD-NoFOG and the other groups were not statistically significant. SNc degeneration is widely regarded as the substrate for parkinsonian motor findings generally, whereas FOG does not usually correlate with the cardinal features of parkinsonism, i.e. tremor, rigidity and bradykinesia.^53–55^ Therefore, it is interesting and perhaps surprising that SNc volume is significantly reduced in PD-FOG when stratifying by FOG status. Reduced SNc volume in PD-FOG supports a major contribution from nigral dopaminergic system degeneration in the mechanism of FOG, and we may speculate that this relates to nigral projections other than the nigrostriatal tract alone, to account for its associations with cognitive impairment (e.g., the nigral-prefrontal projection).^56^

To interpret NM-MRI results in neurodegenerative disease states, the impacts and interactions between aging and the disease must be considered. As shown in **Figure 6A and 6B**, we found that in the combined PD group there is a significant negative correlation between SNc volume and age, whereas no significant correlation exists between these features in older adult controls. The presence of this relationship between age and SNc volume in the PD group, but not the control group suggests that there is an interaction between aging and PD that is strongly associated with SNc neurodegeneration.

This finding remained significant when controlling for disease duration, indicating that this was not simply due to older PD patients with longer disease durations. This is a convergent finding in that it is already known that PD has increasing incidence with aging, and cellular mechanisms for this relation between PD and aging have been identified in model systems, such as increasing mitochondrial dysfunction that occurs with aging.^57, 58^ However, to our knowledge, this is the first report of *in vivo* observation of a significant relationship between aging and SNc degeneration in PD patients.

Importantly, it appears that this relationship is particularly strong in PD patients with > 5 years disease duration (**Figure 6D**). This is again consistent with the clinical observation that patients with older age of onset tend to have a more rapid decline than patients with younger onset PD ^59^. This finding is also consistent with our proposed schema for neuromelanin accumulation and loss in aging and disease, shown in **Figure 1**. As the disease progresses, the period of predominant cell sickness (and complex neuromelanin dynamics) ends, and the period of predominant neurodegeneration ensues with progressive loss of neuromelanin. Also, in keeping with this hypothesis, we found that in PD patients with disease duration > 5 years, LC volume is significantly negatively correlated with age (**Figure 6C**). This is the first *in vivo* report of a significant correlation between LC volume and age in PD to our knowledge, as well.

We observed that in PD-FOG there is a significant negative correlation between SNc volume and ON-medications MDS-UPDRS-III and FOG-Q score. However, in PD-NoFOG these correlations are not seen. These findings are shown in **Figure 5**. FOG is an advanced PD symptom that is associated with falls and cognitive impairment, which are also features of severe disease. The conflicting results in prior studies that did not find any significant correlation between SNc NM-MRI measures and MDS-UPDRS-III may have been due to the inclusion of entirely or almost entirely early to moderate PD and few, if any, PD patients with more severe motor symptoms (e.g., FOG or ON-medications MDS-UPDRS-III > 45). This is again consistent with the hypothesized model in **Figure 1** in that NM-MRI contrast in early or less severe disease is expected to reflect competing effects on neuromelanin due to both cell sickness (neuromelanin increase) and neurodegeneration (neuromelanin loss) and a linear association between motor symptoms and NM-MRI contrast measures in SNc will not be observed. However, when neurodegeneration predominates in more severe disease (i.e., PD-FOG) net neuromelanin loss will also predominate, and a linear correlation between the neurodegenerative phenotype (increasing MDS-UPDRS-III score, increasing FOGQ score) and SNc volume loss can be observed. The results observed in this analysis are consistent with this interpretation. In the analysis stratifying participants based on disease duration, in the PD group with duration > 5 years, a similar association was observed between SNc volume and FOGQ score, but no significant association was seen between SNc volume and MDS-UPDRS-III in this group. This may be because disease duration alone is not a robust indicator of PD severity, given that PD progression is highly heterogeneous. Some patients will retain good levodopa response and stable clinical syndrome for much longer than others.^59, 60^

We found significant positive correlations between SNc volume and global cognition (MoCA) and cognitive reserve (years of education) in the PD-FOG group. No significant correlations between these measures were seen in the PD-NoFOG group. Although SNc neurodegeneration has long been implicated in the motor symptoms of PD, the notion that cognitive impairment in PD is due to rostral spread of synuclein pathology to cortical structures has become popular in the context of the Braak hypothesis ^61^. However, the role of SNc in cognition could not be well assessed prior to the advent of NM-MRI due to lack of an imaging contrast to define the ROI. The results shown in **Figure 4** indicate that SNc volume is implicated in cognition in PD, and they suggest a novel association between cognitive reserve and SNc volume. Again, the identification of these correlations in the PD-FOG group and not in the PD-NoFOG group indicates that linear relationships between NM-MRI measurements of SNc volume and disease phenotype are most readily observed in the more advanced disease stages. A further implication of these results is that traditional localization of PD cognitive impairments, such as “fronto-striatal cognitive impairment” are likely incomplete and do not reflect the network organization of the nigral dopamine system, with wide-ranging projections in many brain regions that are impacted by the health of neurons with cell bodies in SNc. These results warrant further investigation and replication in a larger cohort.

### Limitations and Future Directions

The PD-FOG and PD-NoFOG groups did not all have a complete battery of neuropsychological testing, and only MoCA was available from all participants for assessment of cognition. A larger study is needed with detailed neurocognitive testing on all subjects to confirm the association between cognition and SNc volume assessed with NM-MRI. We are currently recruiting PD-FOG and PD-NoFOG participants and acquiring a detailed neurocognitive battery for this purpose. In this ongoing study we will also perform analyses to further explore the relationship between cognition and motor dysfunction in PD-FOG vs. PD-NoFOG. We also plan recruitment of an age-matched control group for comparisons.

Assessment of the contribution of cerebrovascular disease to cognitive and motor impairment would be useful to add context for the findings shown here. Simultaneous study with both NM-MRI and pulse sequences sensitive to cerebrovascular disease burden would allow this, and we are currently collecting datasets for this purpose in a future study. No MRI dataset suitable for assessment of cerebrovascular disease burden is available for the current study, and this is a limitation. It would also be useful to investigate the associations examined in this work with further stratification of the PD-FOG group based on levodopa response. We did not have enough individuals with FOG characterized based on levodopa response to power such analyses in this study. Finally, although these results support our mechanistic hypothesis, we cannot rigorously test mechanistic hypotheses that involve cell and molecular biology concepts in human studies alone. Additional studies in model systems and radiologic-pathologic correlation studies are needed to further investigate the mechanistic role of neuromelanin in PD.

## Conclusion

NM-MRI is a rapidly emerging neuroimaging modality with established capacity to detect neurodegenerative disease effects, but it remains underutilized to study PD pathogenesis and the neural substrates of PD phenomenology. The cognitive and motor correlations identified in this study enhance our understanding of the role of LC and SNc in PD, but also demonstrate the potential utility of NM-MRI in investigations of their biology in health and aging. The correlations identified between aging and SNc and LC volumes in advancing PD provide the first *in vivo* evidence of the interaction between aging and PD to drive neurodegeneration in melanized catecholamine nuclei in humans.

## Data Availability

All data produced in the present study are available upon reasonable request to the authors.

## Declarations

### Funding

Xiaoping Hu and Daniel Huddleston receive funding from the Michael J. Fox Foundation (MJFF 10854 and MJFF 010556). Lucas McKay and Daniel Huddleston are supported by the American Parkinson’s Disease Association (APDA) Center for Advanced Research at Emory. Lucas McKay is also supported by the McCamish Parkinson’s Disease Innovation Program at Georgia Tech. Stewart Factor and Daniel Huddleston are supported by the Curtis Family and the Sartain Lanier Family Foundation. Dr. Factor is also supported by NIH grant funding (U10 NS077366). Daniel Huddleston is also supported by NIH grant funding (NIH-NINDS 1K23NS105944-01A1; NIH-NINDS 1U19AG071754), the Emory Lewy Body Dementia Association (LBDA) Research Center of Excellence, and the McMahon Family. The Emory MRI facility used in this study is supported in part by funding from a Shared Instrumentation Grant (S10) grant 1S10OD016413-01 to the Emory University Center for Systems Imaging Core. Recruitment of control individuals for this research was facilitated by the Emory Alzheimer’s Disease Research Center (NIH-NINDS P50-AG025688).

### Conflict of interest/Competing interests

Daniel Huddleston is inventor on an issued patent (U.S. patent # 9600881, 3/21/17) which covers aspects of the neuromelanin-sensitive MRI methods discussed here, as well as a patent application (US- 2021/0007603-A1) which covers aspects of the analysis method presented here. Stewart Factor has the following disclosures: Honoraria: Lundbeck, Sunovion, Biogen, Acorda, Alterity, Takeda. Grants: Medtronics, Boston Scientific, Sun Pharmaceuticals Advanced Research Company, Biohaven, Impax, Sunovion Therapeutics, Neurocrine, Vaccinex, Voyager, Addex Pharma S.A., Prilenia Therapeutics CHDI Foundation, Michael J. Fox Foundation, Parkinson Foundation. Royalties: Demos, Blackwell Futura, Springer for textbooks, Uptodate Other Signant Health (Bracket Global LLC) Lucas McKay performs technical consulting for Biocircuit Technologies. Richa Tripathi has following disclosures: Honoraria: UpToDate.

### Availability of data and material

Upon reasonable request to the contact author.

### Code availability

Not applicable.

### Ethics Approval

This research was carried out according to a protocol approved by the Emory Institutional Review Board and in accordance with the Declaration of Helsinki.

## Notes

### Funding Statement

Daniel Huddleston and Xiaoping Hu receive funding from the Michael J. Fox Foundation (MJFF 10854 and MJFF 010556). Daniel Huddleston and Lucas McKay are supported by the American Parkinson Disease Association (APDA) Center for Advanced Research at Emory. Lucas McKay is also supported by the McCamish Parkinson Disease Innovation Program at Georgia Tech. Stewart Factor and Daniel Huddleston are supported by the Curtis Family and the Sartain Lanier Family Foundation. Dr. Factor is also supported by NIH grant funding (U10 NS077366). Daniel Huddleston is also supported by NIH grant funding (NIH-NINDS 1K23NS105944-01A1; NIH-NINDS 1U19AG071754), the Emory Lewy Body Dementia Association (LBDA) Research Center of Excellence, and the McMahon Family. The Emory MRI facility used in this study is supported in part by funding from a Shared Instrumentation Grant (S10) grant 1S10OD016413-01 to the Emory University Center for Systems Imaging Core. Recruitment of control individuals for this research was facilitated by the Emory Alzheimer Disease Research Center (NIH-NINDS P50-AG025688).

### Author Declarations

The Institutional Review Board of Emory University gave ethical approval for this work.

